# Integrated transcriptomics and proteomics analysis reveal age-related prognosis and myocardial fibrosis in pediatric DCM

**DOI:** 10.1101/2025.09.26.25336775

**Authors:** Yan Zhang, Zubo Wu, Lin Wang, Xihang Fu, Jing Wu, Xiaodie Li, Jie Liu, Peng Zhu, Nianguo Dong, Jiawei Shi, Hua Peng

**Author notes:** These authors correspondence to this work **Corresponding Author:** Hua Peng, Department of Pediatric, Union Hospital, Tongji Medical College, Huazhong University of Science and Technology, Wuhan, Hubei, 430030, China., Jiawei Shi, Department of cardiovascular surgery, Union Hospital, Tongji Medical College, Huazhong University of Science and Technology, Wuhan, Hubei, 430030, China. These authors contributed equally to this work (Yan Zhang, Zubo Wu, Lin Wang).

## Abstract

**Background:** Dilated cardiomyopathy (DCM) represents the most prevalent form of pediatric cardiomyopathy. However, the characteristics and etiology of this condition across different pediatric age groups remain incompletely elucidated.

**Methods and results:** Data from 88 pediatric patients aged <14 years diagnosed with primary DCM between January 2021 and December 2023 were retrospectively retrieved from the institutional database and analyzed. Patients were stratified into two groups using a 3-year age cutoff. Clinical characteristics, echocardiographic parameters, and biomarkers were documented for each patient to facilitate subsequent analysis. Of these patients, 11 underwent heart transplantation and additional transcriptomic and proteomic analyses were conducted to identify differentially expressed genes (DEGs) and proteins (DEPs). Additionally, cardiac magnetic resonance imaging was utilized to evaluate myocardial fibrosis. Statistically significant disparities were observed between the two age groups with respect to clinical characteristics and clinical outcomes. Patients in the ≤3-year-old group exhibited a more favorable prognosis. Multi-omics integrative analysis revealed that genes associated with inflammatory responses and energy metabolism impairment—notably *PDK4, CHI3L1, ZBP1* and *ANXA1*—were enriched in the younger group. In contrast, myocardial inflammation and fibrosis were the predominant features in the older group, as evidenced by the significant upregulation of *COL1α1, CTGF, RO60* and *MAT2B*.

**Conclusions:** At both the transcriptomic and translational levels, we uncovered global variations in miRNAs and proteins associated with DCM, following stratification of patients by the 3-year age cutoff. Specifically, energy metabolism dysregulation plays a pivotal role in younger pediatric patients, whereas irreversible myocardial fibrosis may underlie the suboptimal response to conservative therapy in older pediatric patients.

## Introduction

Dilated cardiomyopathy (DCM) refers to structural (ventricular or biventricular dilatation) and functional (impaired contractility) abnormalities caused by genetic mutations, inflammation, autoimmunity and exposure to chemicals and toxins[1]. Cardiac muscle disorders may lead to considerable morbidity and mortality due to malignant complications[2].Early diagnosis of DCM in children is challenging due to its atypical early symptoms such as poor feeding, respiratory distress and growth failure[3, 4]. Over the past few decades, the prognosis of patients with DCM has shown limited improvement, largely attributable to conventional management predicated on the foundational therapies for reduced ejection fraction (EF)[5]. Nearly half of pediatric patients die or undergo cardiac transplantation within 5 years of initial diagnosis, the rest may achieve a certain degree of cardiac function recovery through conservative medical treatment[6].

Abnormal transcriptional and functional alterations in multiple cardiac cell types, such as cardiomyocytes, fibroblasts and immune cells, constitute a pathological mechanism of DCM hearts[7]. RNA sequencing stands as the most robust tool for high-throughput quantitative transcriptomics, enabling the accurate quantification of gene activity—even with minimal tissue volumes. Furthermore, proteomic technologies provide an alternative strategy for conducting comprehensive, unbiased analyses of protein expression and post-translational modifications[8]. Notably, the combination of RNA sequencing and proteomics facilitates a more thorough understanding of pathogenesis and disease progression.

In this study, we seek to preliminarily investigate the pathogenic mechanisms underlying dilated cardiomyopathy (DCM) in pediatric patients across different age groups, through comparative analyses of clinical characteristics integrated with RNA sequencing and proteomic data. These endeavors are designed to lay a theoretical basis for the development of therapeutic strategies for DCM.

## Methods

### Study participants

A total of 88 unrelated pediatric patients with DCM of Han Chinese ethnicity were recruited from Wuhan Union Hospital from 2021 to 2023. Patients were stratified into two groups: young children (< 3 years) and older ones (≥ 3 years). Clinical characteristics of these patients were collected from medical records. Inclusion criteria were defined based on echocardiographic findings at diagnosis[9]. Furthermore, patients were excluded if they were presented with: (1) secondary DCM caused by developmental abnormality such as myocardial ischemia secondary to abnormal vascular development; or (2) DCM with a mixed phenotype (e.g., those displaying restrictive or hypertrophic features). Among these DCM patients, 11 underwent heart transplantation owing to severe disease complications during subsequent treatment, with 2 aged < 3 years and the remaining 9 aged ≥ 3 years. Heart tissue samples from the left atria and left ventricle (LV) of all DCM patients were obtained following heart transplantation process. These samples were immediately flash-frozen and then stored at -80 °C until later use. Meanwhile, flash-frozen heart tissue samples from donors were obtained from myocardial biopsies of heart donors prior to HT. The utilization of tissues for research purposes was reviewed and approved by the Institutional Review Boards of both the Gift-of-Life Donor Program and Wuhan Union Hospital, with the review process strictly adhering to the ethical guidelines outlined in the Declaration of Helsinki. Prior to the initiation of the study, written informed consent was formally obtained from the legal guardians of each participating individual to ensure compliance with ethical standards for human research.

### RNA extraction and sequencing

Total RNA was extracted from cardiac tissues using Trizol Reagent (Invitrogen Life Technologies). RNA purity and concentration were assessed with a NanoDrop spectrophotometer (Thermo Scientific) and Qubit 4.0, while RNA integrity and quantity were evaluated using the Agilent 2100/4200 system. In the process of library construction, a minimum of 1 μg total RNA per isolate was employed. For this step, Hieff NGS® DNA Selection Beads were used for purification and fragment selection, and polymerase chain reaction was carried out for amplification and enrichment. Ultimately, RNA sequencing was performed on the DNBSEQ-T7 platform employing combinatorial probe-anchor synthesis.

### Protein extraction and tag label-free mass spectrometry analysis

Heart tissue samples (100 mg) were homogenized using an MP FastPrep-24 homogenizer (24×2 format, 6.0 m/s, 60 seconds, two cycles) in lysis buffer consisting of 2.5% SDS/100 mM Tris-HCL (pH 8.0). Following centrifugation, proteins present in the supernatant were precipitated via the addition of 4 times of ice-cold acetone. Subsequently, after undergoing sequential steps including the dissolution of the protein pellet, reduction reaction, and alkylation reaction, the protein concentration was determined using the Bradford method. The peptide eluate was then vacuum-dried and stored at -20℃ for subsequent use.

Liquid chromatography-tandem mass spectrometry (LC-MS/MS) was conducted on a Q Exactive Plus mass spectrometer coupled with an Easy-nLC 1200 system to detect protein content. Finally, raw LC-MS/MS data were analyzed using MaxQuant (v1.6.6) with the Andromeda database search algorithm, and spectral files were queried against the UniProt Human proteome database.

### Cardiac magnetic resonance imaging

After excluding 10 children who were deceased or lost to follow-up, 77 remaining patients returned to the hospital for further treatment. Participants underwent a standardized imaging protocol on a 3T scanner (MAGNETOM Skyra system; Siemens Healthcare, Erlangen, Germany) equipped with an 8-channel phased-array body coil. Following gadolinium administration, steady-state free precession cine images were obtained in contiguous short-axis slices spanning from the basal to apical levels, as well as in three long-axis views (2-chamber, 3-chamber, and 4-chamber). Key scanning parameters were set as follows: repetition time (TR): 3.42 ms; echo time (TE): 1.23 ms; flip angle: 60°; field of view (FOV): 250 × 300 mm^2^; matrix size: 162 × 192; average temporal resolution: 35–45 ms; and slice thickness: 8 mm.

### Statistical Analysis

Variables were summarized as mean ± standard deviation (SD), median with interquartile range (IQR), or number (percentages), as appropriate. Continuous variables were expressed as medians and interquartile ranges, and intergroup differences were compared using the Wilcoxon rank‐sum test. Categorical variables were presented as number (percentages) and analyzed with unpaired, two-tailed student’s t-test. GraphPad Prism 8 software was used for statistical analyses. Differences were considered statistically significant at *P* < 0.05.

Differentially expressed genes (DEGs) were filtered using the “DESeq2” R package. Thresholds for defining upregulated and downregulated DEGs were set as log_2_-transformed fold change (log_2_FC) > 1 or log_2_FC < -1, respectively, combined with a corrected false discovery rate (FDR) of < 0.05. Thereafter, proteins overlapping with the identified DEGs were extracted from the proteomic profiling dataset. For this subset of proteomic data, the relative abundance of each protein was first subjected to log2 transformation, and Student’s t-test was employed to evaluate proteins with significant expression differences between the two groups. Differentially expressed proteins (DEPs) were classified as significantly upregulated if they met FDR-corrected *P* < 0.05 and fold change (FC) > 1.5, and significantly downregulated if they satisfied FDR-corrected *P* < 0.05 and FC < 0.67.

## Results

### Impacts of age on clinical symptoms and outcomes

Table 1 presented a comparison of characteristics between newly diagnosed young children and older ones with DCM. In our study, 43 of the patients (48.9 %) were males. In total, 11 patients (12.5 %) clinically presented with shock and 7 (7.9 %) had ascites or embolism at initial diagnosis. Hepatomegaly was observed in 44 patients, with a significantly higher prevalence among children aged ≥ 3 years. For the younger group, 22 children (53.7 %) demonstrated a more favorable prognosis, as assessed by ejection fraction (stabilizing at over 50 % or increasing by more than 10 % following six months of standardized drug therapy). Furthermore, younger patients exhibited lower NYHA classification, less LVDD together with valvular regurgitation at diagnosis. Obviously aortic valve regurgitation was observed in both groups. Specifically, mitral and tricuspid regurgitation were more common in older children than in younger ones. Aortic regurgitation was rare, with no pulmonary regurgitation detected (data not shown). Notably elevated serum Cr was also identified in DCM patients aged < 3 years.

**Table 1.** Comparisons of clinical characteristics between pediatric patients with initial diagnostic age <3 years and ≥3 years.

| | All<br>(n = 88) | <3 years<br>(n = 41) | $\geq 3$ years<br>(n = 47) | p-value |
| --- | --- | --- | --- | --- |
| Male, n (%) | 43(48.9) | 18(43.9) | 25(53.2) | 0.385 |
| **Main presenting symptoms** |  |  |  |  |
| Shock, n (%) | 11(12.5) | 6(14.6) | 5(10.6) | 0.572 |
| Hepatomegaly, n (%) | 44(50.0) | 15(36.6) | 29(61.7) | 0.019 |
| Ascites, n (%) | 7(7.9) | 2(4.8) | 5(10.6) | 0.063 |
| Embolism, n (%) | 7(7.9) | 5(12.1) | 2(4.3) | 0.227 |
| Favorable prognosis, n (%) | 31(35.2) | 22(53.7) | 9(19.1) | 0.001 |
| NYHA classification, median<br>(25th-75th percentile) |  | 2(2,4) | 4(3,4) | 0.011 |
| **Echocardiography parameters** |  |  |  |  |
| LVDD (mean ± SD) | | 41.88±10.25 | 51.23±8.64 | <0.001 |
| LVEF, (%) |  | 27(20,33) | 24.4(20,30) | 0.27 |
| LVNC, n (%) | 44(50.0) | 16(39.0) | 28(59.6) | 0.544 |
| Pericardial effusion, n (%) | 37(84.1) | 17(41.5) | 20(42.6) | 0.918 |
| Valvular regurgitation, n (%) | 84(95.5) | 37(90.2) | 47(1.00) | 0.043 |
| MR, median (25th-75th percentile) |  | 1(0,2) | 1(1,3) | 0.037 |
| TR, median (25th-75th percentile) |  | 1(0,1) | 1(1,2) | 0.034 |
| AVR, median (25th-75th percentile) |  | 0(0,1) | 0(0,0) | 0.324 |
| **Electrocardiographic abnormalities, n (%)** | 69(78.4) | 33(80.5) | 36(76.6) | 0.658 |

LVDD, left ventricular diastolic dysfunction; LVEF, left ventricular ejection fraction; LVNC, left ventricular noncompaction; MR, mitral regurgitation; TR, tricuspid regurgitation; AVR, aortic valve regurgitation

**Table 2.**
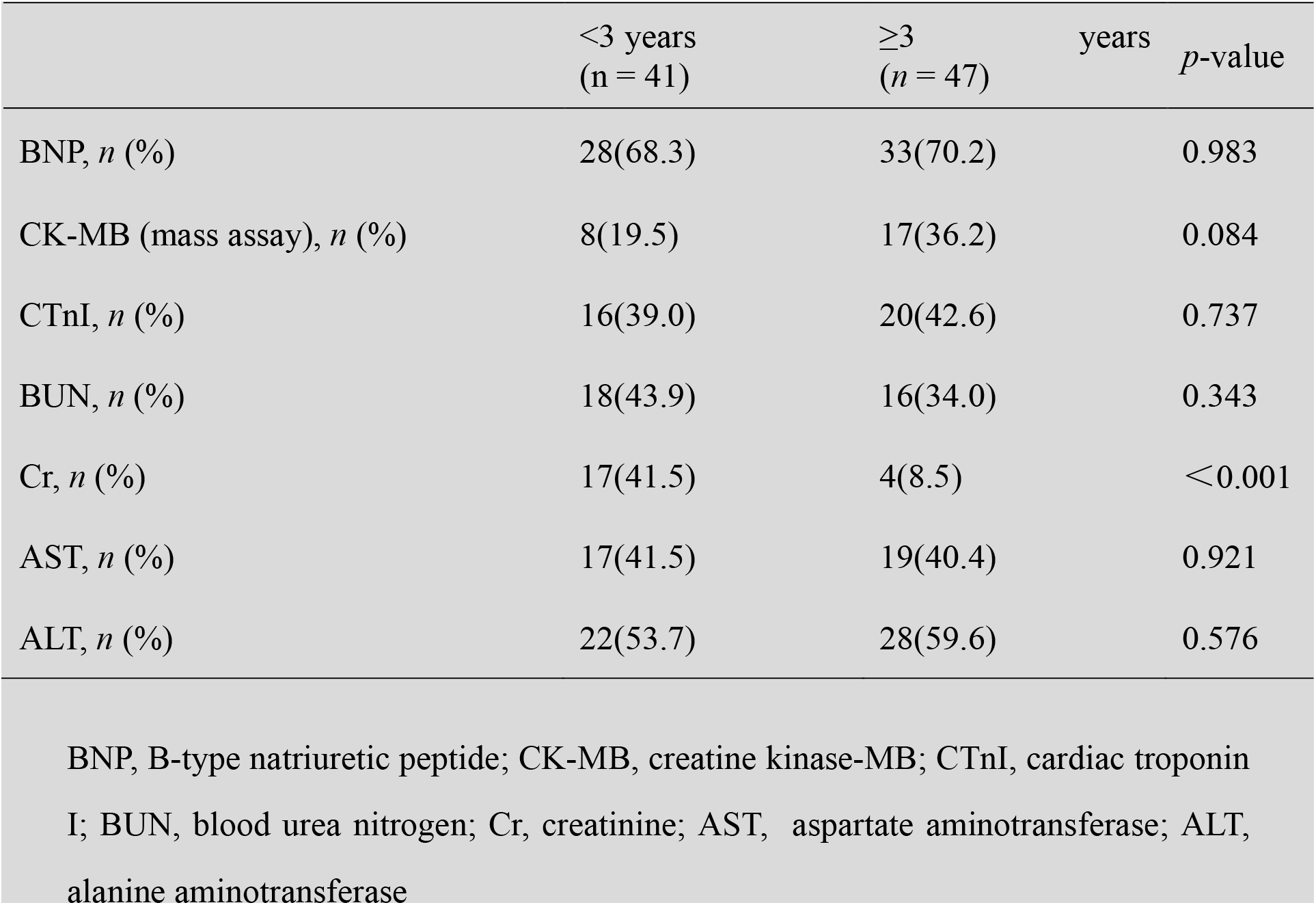
Analysis of serological markers in children with DCM at initial diagnosis.

**Table 3.**
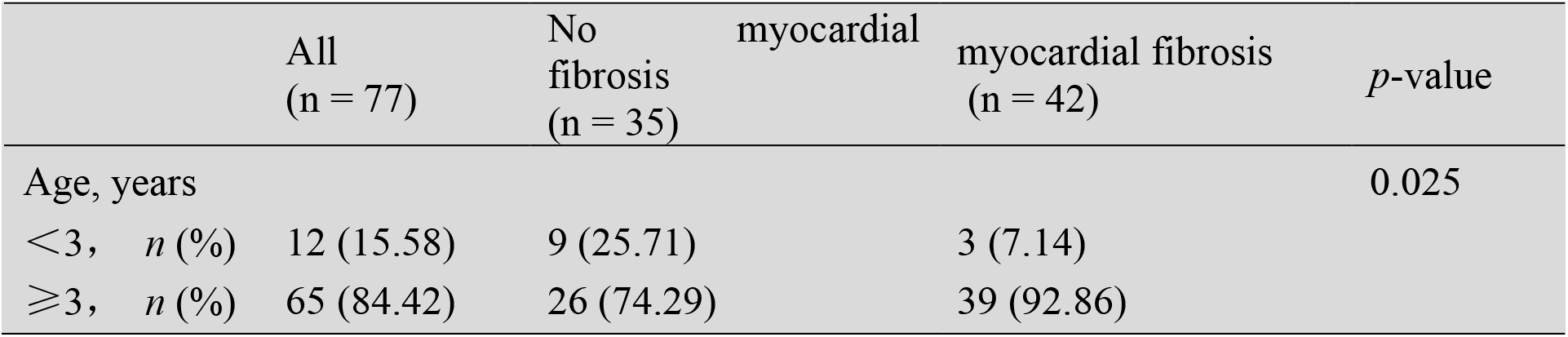
Cardiac MRI findings in pediatric patients with DCM.

### Identification of differential genes

A total of 11 DCM patients underwent heart transplantation, and their clinical characteristics are presented in Table S1. Using 3 years of age as the cutoff, these patients were separated into the younger HT group and the older HT one. Following a stringent QC procedure, 17,320 genes were detected in the present transcriptional analysis. Using a filtering threshold of|log_2_FC|>1 together with adjusted *P* < 0.05, 60 upregulated and 96 downregulated transcripts were identified in the comparison between the two groups (Figure 1A).

**Figure 1.**
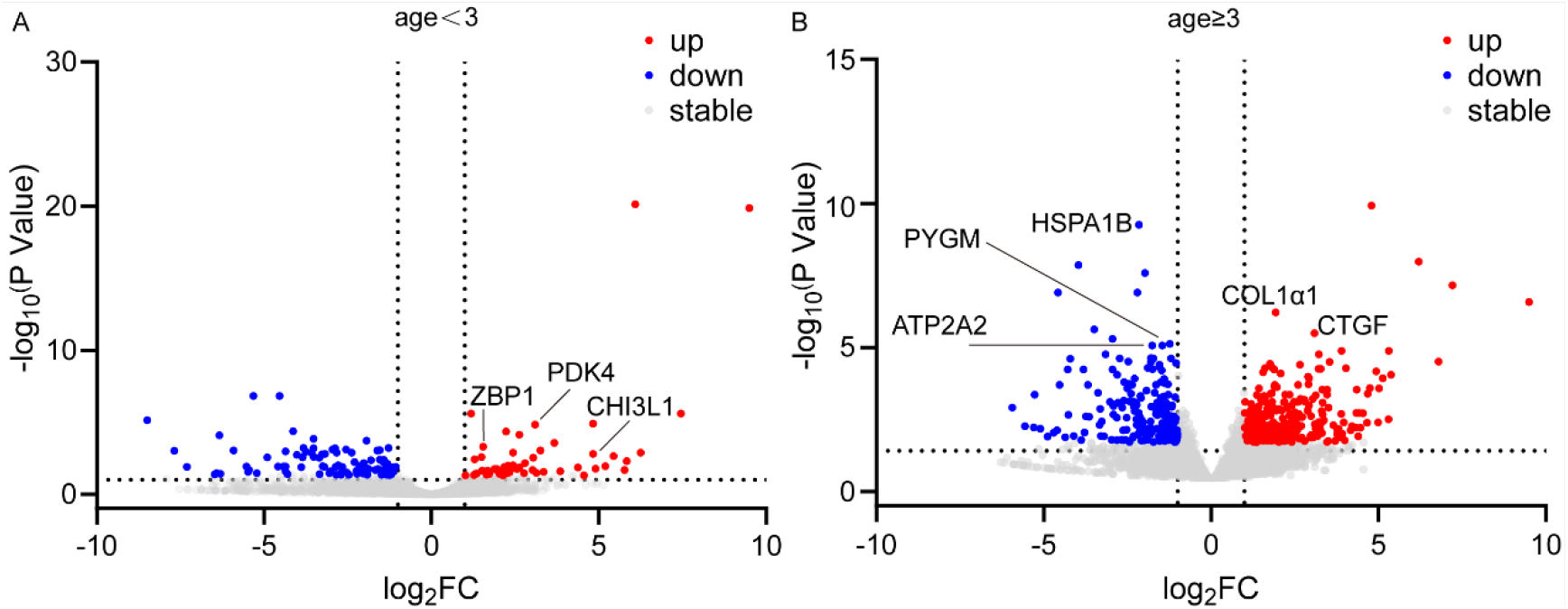
Identification of global differences in mRNAs. Volcano plot of differentially mRNAs between (**A**) younger DCM patients and healthy control and (**B**) older DCM patients and healthy control. Red, green, and grey colored dots indicated up-regulation, down-regulation and no significant change in transcriptomic levels in the former group relative to the latter one.

After the rigorous QC process, we identified 156 differentially expressed genes between younger patients and the control subjects, including 60 upregulated and 96 downregulated transcripts, which can act as potential candidates for further investigation into novel biomarkers for diagnosing DCM in early childhood. It is noteworthy that previously well-characterized glucose oxidation biomarkers implicated in the regulation of glucose oxidation, such as pyruvate dehydrogenase kinase 4 (*PDK4*) were encompassed within upregulated genes identified in younger DCM patients compared to healthy controls[10]. Inflammatory responses may also contribute substantially to the development of DCM, given that the expression levels of Chitinase-3 like-protein-1 (*CHI3L1*) and Z-DNA binding protein 1 (*ZBP1*) were markedly increased as well (Figure 2A). In patients over 3 years of age, 286 genes were increased, and 214 genes were downregulated. Among these, genes related to energy metabolism—specifically ATPase sarcoplasmic/endoplasmic reticulum Ca^2+^ transporting 2 (*ATP2A2*), heat shock 70 kDa protein 1B (*HSPA1B*), and muscle glycogen phosphorylase (*PYGM*)—exhibited significantly decreased expression. In contrast, genes associated with myocardial fibrosis, such as collagen type I alpha 1 chain (*COL1α1*) and connective tissue growth factor (*CTGF*), showed a marked upregulation in expression (Figure 2B).

**Figure 2.**
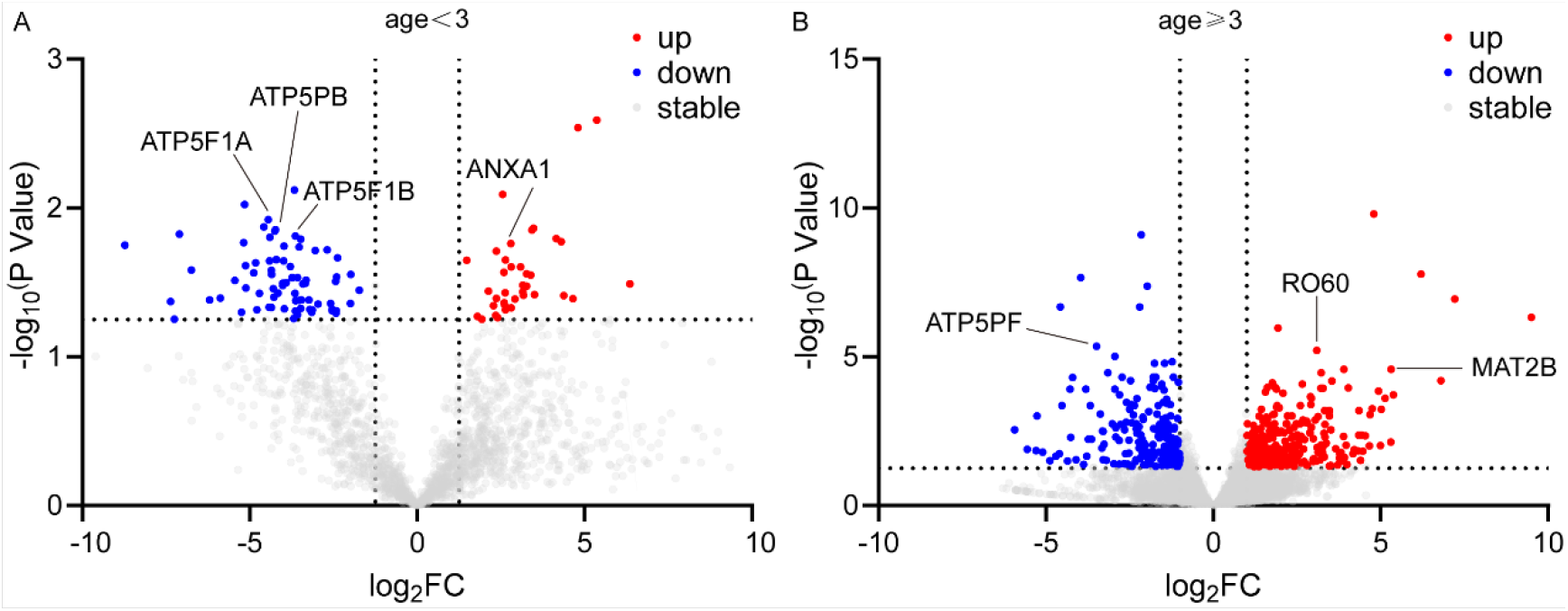
Differential proteins in DCM patients. Volcano plot of proteins between (**A**) younger DCM patients and healthy control and (**B**) older DCM patients and healthy control.

### Proteomics analyses between DCM and normal myocardium

To elucidate the underlying mechanisms of DCM, we further performed quantitative proteomic analyses on DCM samples. In patients aged < 3 years, isobaric tags for relative and absolute quantification (iTRAQ) analysis identified 37 upregulated proteins and 75 downregulated proteins. Consistent with our transcriptomic data, the inflammatory response was enriched in younger patients, accompanied by increased expression of annexin A1 (ANXA1). Impairments in myocardial energy metabolism were observed, potentially attributed to the downregulated expression of multiple ATPase enzymes. For older patients, the proportion of energy metabolism disorders appeared to decrease, with the main manifestations concentrating on myocardial inflammation and fibrosis, as evidenced by the increased expression of RO60 and beta subunit of methionine adenosyltransferase (MAT2B).

### MRI examination

Cardiac MRI was performed in patients with a confirmed DCM diagnosis at a later stage (6 months after diagnosis) to further characterize the pathological features of DCM. Among 12 pediatric DCM patients aged < 3 years, only 3 had significant myocardial fibrosis, mainly localized to the left ventricular apical segment, left ventricular subendocardial layer and left ventricular mid-myocardial layer. In patients aged ≥ 3 years, a majority (39 of 65 patients) showed evidence of myocardial fibrosis, predominantly distributed in the left ventricular septal wall, left ventricular subendocardial layer, and left ventricular mid-myocardial layer; in some cases, both the left and right ventricles were involved. This finding suggests that myocardial fibrosis may constitute a primary pathogenic factor in DCM in older children.

## Discussion

In the present study, we examined the clinical characteristics and disease outcomes of pediatric DCM stratified by the age of 3 years. Our findings demonstrated that younger patients were more likely to achieve favorable prognosis following standardized pharmacotherapy. By means of multi-omics integration, a catalog of mRNAs and proteins in younger and older patients were identified respectively. By comparing the differences, we speculated that DCM in younger children is predominantly driven by infections and abnormal energy metabolism, which tend to yield favorable prognoses following anti-infective therapy and myocardial nutritional support. However, irreversible myocardial fibrosis plays a crucial role in DCM among older children.

Research on pediatric DCM remains limited; treatment strategies for these patients are often extrapolated from adult therapeutic evidence, with a paucity of studies investigating age-related characteristics, disease severity, and clinical outcomes. Studies in adult populations have demonstrated that the incidence of DCM increases with age, predominantly affecting middle-aged and elderly individuals. It is typically diagnosed in individuals aged between 20 and 50[11]. Most patients with cardiomyopathy were asymptomatic in early stages and progressively worsened with age[12]. Long time hemodynamic changes resulted in heart failure manifestations such as feeding difficulties, dyspnea, tachypnoea, hepatomegaly, and oedema, due to hemodynamic changes[13]. Nearly 49 % of children underwent heart transplantation within 5 years of diagnosis. Congestive heart failure, an increased left ventricular end-systolic dimension z-score, and older age at diagnosis were identified as factors that elevate the risk of death or heart transplantation[14, 15]. Similarly, in our study, older children with DCM were more frequently presented with manifestation of congestive heart failure hepatomegaly and an increased LVDD at initial diagnosis; their treatment response was inferior to that of younger children.

Numerous biomarkers have been proposed as outcome indicators of DCM. A study on adult DCM demonstrated that higher levels of Cr were positively correlated with mortality in patients[16]. Elevated levels of osteopontin (OPN), a circulating fibrotic biomarker, were associated with increased serum Cr levels, suggesting that renal dysfunction is associated with a greater fibrotic burden[17]. Our results demonstrated that elevated Cr levels were more pronounced in younger DCM patients. This may be attributed to the higher susceptibility of younger DCM children to concurrent renal impairment, which is gradually ameliorated as treatment proceeds.

Heterogeneous aetiologies, such as gene mutations, chronic cardiac inflammation and abnormal energy metabolism ultimately result in DCM, with a histological hallmark of cardiac fibrosis[2]. After bacterial and viral invasion or autoimmunity, inflammation process triggered, and immune cells accumulate to the heart to repair the myocardium[18]. Cytokines released from immune cells promote remodeling, collagen deposition and fibrosis[19, 20]. In our study, pediatric patients of different age groups demonstrated genetic alterations linked to myocardial fibrosis and abnormal energy metabolism. Younger age group was primarily affected by abnormal energy metabolism, as evidenced by the increased expression of PDK4—a protein that has been confirmed to regulate the oxidation of glycolytically derived pyruvate in cardiomyocytes[10, 21]. Inflammation responses also exerted a pivotal role in this process, since *CHI3L1, ZBP1* and *ANXA1* were confirmed to be closely associated with myocardial inflammation[22-24]. Whereas patients with DCM aged ≥ 3 years exhibited a higher incidence of myocardial fibrosis, as evidenced by the increased expression of fibrosis-related genes *COL1α1* and *CTGF*, which was further validated by MRI analysis. This is consistent with the clinical observation that young children exhibit better therapeutic efficacy with anti-infective and myocardial nutritional therapies, while older children tend to require heart transplantation[25-27].

However, there are several limitations in this study. Our study was a retrospective study from a single institution with a limited sample size. Following the identification of target genes or proteins via multi-omics integration, it is advisable to further validate them through *in vivo* and *in vitro* experiments, which constitutes our subsequent work. Besides, more detailed serological results should be collected to further assess the prognosis of pediatric patients with DCM.

In summary, we identified global differences in miRNAs and proteins associated with DCM, stratified by age at 3 years, at the transcriptomic and translational levels. Many of them have been demonstrated to be classical biomarkers. Energy metabolism abnormalities play a crucial role in younger patients and irreversible cardiac fibrosis may account for the poor therapeutic efficacy of conservative treatment in older pediatric patients.

## Data Availability

The data are available from the corresponding author on reasonable request.

## Acknowledgements

This work was supported by National Key R&D Program of China (No.2023YFC2706200), Wuhan Knowledge Innovation Special Project (No.2023020201010160), key research and development program by Hubei province of China (No.2022BCA043), Natural Science Foundation of Hubei Province of China (No.2020CFB813, No.2020CFB764) and the National Natural Science Foundation of China (No. 82200990).

